# Long-Term Forecasting of a Motor Outcome Following Rehabilitation in Chronic Stroke via a Hierarchical Bayesian Model of Motor Learning

**DOI:** 10.1101/2022.10.20.22280926

**Authors:** Nicolas Schweighofer, Dongze Ye, Haipeng Luo, David Z. D’Argenio, Carolee Winstein

**Affiliations:** Biokinesiology and Physical Therapy, University of Southern California, Los Angeles; Computer Science, University of Southern California, Los Angeles; Biomedical Engineering, University of Southern California, Los Angeles

**Keywords:** Chronic stroke, neurorehabilitation, motor learning, forecasting model, hierarchical Bayesian modeling, Dynamical models

## Abstract

**Background:** Given the heterogeneity of stroke, it is important to determine the best course of motor therapy for each patient, i.e., to personalize rehabilitation based on predictions of long-term outcomes. Here, we propose a Hierarchical Bayesian dynamical (i.e., state-space) model of motor learning to forecast long-term changes in a motor outcome due to rehabilitation in the chronic phase post-stroke.

**Methods:** The model incorporates the effects of clinician-supervised training, self-training, and forgetting. In addition, to improve forecasting early in rehabilitation, when data are sparse or unavailable, we use a hierarchical Bayesian structure, which incorporates prior information from similar patients. We use this dynamical model to re-analyze Motor Activity Log (MAL) data of participants with chronic stroke included in two clinical trials: 1) the DOSE trial, in which participants were assigned to a 0, 15, 30, or 60-hour dose condition (data of 40 participants analyzed), and 2) the EXCITE trial, in which participants were assigned a 60-hour dose, in either an immediate or a delayed condition (95 participants analyzed).

**Results:** For both datasets, the dynamical model accounts well for individual trajectory in the MAL during and outside of training and better fits the data than other simpler models without the effects of either supervised training, self-training or forgetting or (static) regression models. We then show how the model can be used to forecast the MAL of new participants up to 8 months ahead and how the hierarchical structure improves the accuracy of the predictions early in training when data are sparse. Finally, we verify that this model, despite its simplicity, can reproduce previous findings of the DOSE trial on the efficiency, efficacy, and retention of motor therapy.

**Conclusion:** In future work, such forecasting models can be simulated for different stages of recovery, dosages, and training schedules to optimize rehabilitation for each person.

## Introduction

Recent modeling work has sought to predict the long-term spontaneous recovery of individuals post-stroke from baseline clinical or neural data, e.g., ^1-4^. Whereas such predictions are useful for clinical and research stratification, the neurorehabilitation clinician needs to accurately predict the long-term changes in motor outcomes in response to specific treatments. Using these predictions, the clinician could then determine the best course of motor therapy for each patient, i.e., personalize rehabilitation^5^.

A difficulty is that stroke is heterogeneous and exhibits considerable variability, including in response to motor therapy^6^. It is known that the integrity of the corticospinal tract predicts gains in functional outcomes due to rehabilitation, e.g., ^4,7,8^. However, multiple other factors are also likely to affect these gains, as well as the retention of these gains following rehabilitation. For instance, we have previously shown that the integrity of visuospatial working memory modulated the effect of blocked, but not distributed, training schedules in chronic stroke^9^. In addition, in re-analyses of data of the EXCITE ^10^ and DOSE ^11^ trials, we have shown that approximately one-fourth of participants continued to see improvements in upper extremity (UE) function following training; conversely, another fourth lost most gains in UE function that resulted from therapy^12,13^.

Given this variability in response to therapy, we need a paradigm shift in predictive modeling in neurorehabilitation that, in addition to clinical and lesion data, incorporates repeated measurements of motor outcomes obtained during motor therapy, whenever these measurements become available. Predictive models that primarily consider such repeated measurements indexed in time order (i.e., time-series) are called *forecasting models*. For example, a recent model can more accurately forecast spontaneous recovery 6 months post-stroke when incorporating repeated measurements than only baseline data ^14^. Here, we extend such an approach to forecasting the effect of rehabilitation in chronic stroke.

What should be the form of such forecasting models in neurorehabilitation? Since neurorehabilitation is based on the premise that sensorimotor activity improves motor recovery via brain plasticity, i.e., “changeability”, the models need to account for the changes in outcomes both during movement therapy, when an increase in performance is expected, and outside of therapy, when either a decrease in performance due to forgetting or an increase in performance are possible. Previously, we proposed a piece-wise linear model of changes in a motor outcome in the DOSE clinical trial, in which the periods of therapy marked the limit between the different linear segments^13^. Although this model well accounted for positive and negative changes both during and following therapy, a model of this type cannot generalize to other datasets because it depends on the timing of training and measurements.

We propose a state-space model approach to predict motor outcomes during and following rehabilitation post-stroke. The model has a compact representation and an adjustable time resolution, allowing generalization to different data sets and even to different schedules of therapy for individual patients. The model extends a previous non-linear, first-order state-space model that explained the long-term changes, and the variability in these changes, in arm use following training in the EXCITE trial^12^. This previous model uses a forgetting term to account for the performance decay often observed post-training, at least in subgroups of patients ^12,13^ and a “self-training” term to account for the change in spontaneous use of the paretic limb outside of training when UE function is above a threshold post-therapy ^12,13,15,16^, which further increases future use and function. In the present model, we further account for the response to therapy via an input term proportional to the dose of motor training, as in our previous piece-wise motor learning model ^13^. Indeed, animal studies, meta-analyses, and recent clinical trials with large doses, including the DOSE trial, showed that large training doses improve UE function, e.g., ^11,17-19^.

Previous models in neurorehabilitation typically predict the mean of the future outcome. However, such point estimation of a future outcome is insufficient for clinical decision-making in neurorehabilitation, because clinicians need to account for the uncertainty of the forecast when assessing different treatment options^a^. To provide interval estimation, we utilize the Bayesian approach, which extends our previous work^12^, as Bayesian models naturally deal with uncertainties by focusing on the probability distributions of *all* parameters.

A final difficulty for accurate long-term forecasting in neurorehabilitation, however, is that for each new patient, there is initially no or little data on the effect of motor therapy. A hierarchical Bayesian model^20^ can, in theory, refine the initial predictions by incorporating prior information from similar patients, via “hyper-parameters.” Crucially, these hyper-parameters can be used as individual prior parameters when predicting the response of a new individual when little outcome data are available, i.e., early in therapy.

Here, we therefore propose and test a novel hierarchical Bayesian dynamical modeling framework that can accurately forecast a clinical measure following rehabilitation in chronic stroke. As a testbed of our model, we use the Motor Activity Log (MAL) data from both the DOSE trial ^11^ and the EXCITE trial^10^. We test whether a minimal model with three terms, accounting for forgetting, response to external training, and self-learning, respectively, can better predict the MAL than reduced dynamical models and non-dynamical regression models for these two datasets. Then, using the DOSE data, we simulate the model to forecast the MAL of “new” patients up to 8 months ahead and study the change in the long-term accuracy of the forecasts as additional training data becomes available. We compare the prediction accuracy for models with and without a hierarchical Bayesian structure for different ranges of forecasting. Finally, we validate the model by testing whether it can account for our previous results on the DOSE dataset on the efficacy, efficiency, and retention of motor training in chronic stroke.

## Methods

### Participants and data

We first developed and validated the model with the MAL data of 40 participants enrolled in the DOSE trial conducted at the University of Southern Califrnia ^11^. The participants had mild-to-moderate upper extremity motor impairment chronically after stroke (onset at least five-month before inclusion). Participants were randomly assigned to the 0, 15, 30, and 60-hour doses, with the dosages distributed over three week-long training bouts, separated by one month. The intervention was based on the Accelerated Skill Acquisition Program (ASAP)^21^, which includes elements of skill acquisition through challenging and progressive task practice. The MAL, which measures the participant’s perception of the amount and quality of functional motor tasks by asking them to recall and rate the quality of movement of the paretic arm for 28 activities of daily living ^22^, was collected by blinded assessors in 14 longitudinal assessments administered: 1) twice before the first training bout; 2) immediately before and after each of the three 1-week training bouts; and 3) monthly for six months following the last training bout.

We further validated the model with the data from the EXCITE trial conducted at 7 US sites, in which participants who had a first stroke within the previous 3 to 9 months and with mild-to-moderate impairment were randomly assigned to either an immediate or a delayed Constraint Induced Movement Therapy (CIMT) group ^10,23^. The immediate group received 10 days of therapy from inclusion; the delayed group received 10 days of therapy after a one-year delay. Participants were tested with the MAL, recorded by blinded assessors, immediately before and after the scheduled therapy for both groups, and at 4 months, 8 months, 16 months, 20 months, and 24 months, with a maximum of 9 data points were available. Due to the limited data per person, we excluded participants with missing observations, resulting in 50 and 45 participants in the immediate and delayed groups, respectively.

### The hierarchical Bayesian motor learning model

We developed a hierarchical Bayesian dynamical model of the changes in the MAL both during training and outside of training post-stroke. The model has three levels: an individual measurement level (level 1), a subject level (level 2), and a population level (level 3).

### Level 1. Modeling intra-individual variations (over time)

In level 1, we modeled how the MAL is updated at each time *t* for each subject *i*. The model contains a single state-space equation that updates a motor memory, a sigmoidal non-linearity, and a measurement equation that yields the MAL at time *t*. The state equation updates the latent variables: memory 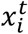 and noiseless, hidden MAL 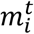as follows:

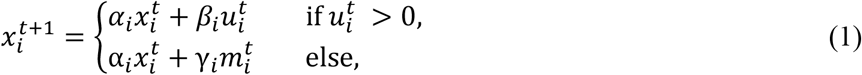

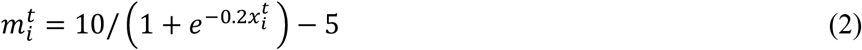

where 0 ≤ *α*_*i*_ ≤ 1 is a forgetting parameter, *β*_*i*_ ≥ 0 a learning rate parameter, *γ*_*i*_ ≥ 0 controls the strength of self-training, and the predicted MAL 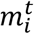ranges from 0 to 5 (the minimum and maximum MAL, respectively) by passing the motor memory through a sigmoidal function. In the DOSE trial, 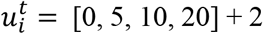is one of the four doses for each of the three weeks of training, including two additional hours of movement testing ^13^ (as a reminder, the total nominal doses are 0, 15, 30, and 60 hours). In EXCITE, 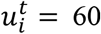, given upon inclusion (immediate group) or 1 year later (delayed group). The constant sigmoid slope of 0.2 was found in preliminary model fitting. Note that no process (state) noise was included, because the data are scarce ^24^.

To account for outliers, the observed MAL 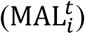 is then modeled with a generalized Student’s *t*-distribution; that is

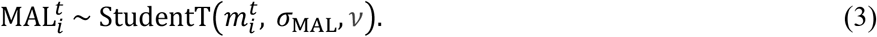

where 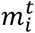(the predicted MAL) is the center of the distribution, *σ*_MAL_ is a scale parameter, and *v* is the degree of freedom. Note that *σ*_MAL_ is subject-independent because we assume the measurement noise is an inherent property of the MAL.

### Level 2. Modeling inter-individual variations

In level 2, we model the individual parameters α_*i*_, β_*i*_ , γ_*i*_, as well as the initial state 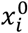withthe following prior distributions:

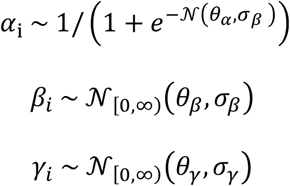

where *N*(*μ σ*) is the normal distribution with mean μ and standard deviation *σ*, the forgetting rate α_*i*_ is constrained between 0 and 1 via the sigmoid function, and the learning rates *β*_*i*_ and self-training rates γ_*i*_ are non-negative with truncated normal distributions (*N*_[0,∞)_). As in our previous model ^13^, the baseline MAL_ini_ is a (linear) co-variate of the initial memory state:

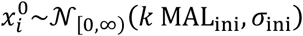

### Level 3. Modeling the population level

In level 3, the hyper-parameters, *θ*_α_, *θ*_β_, *θ*_γγ_, σ_α_, σ_β_, *σ*_γ_, *σ*_ini_, *σ*_MAL_, *k*, and *v* are initially sampled from weakly-informative prior distributions as shown in Table S1. We use normal priors for the location hyper-parameters *θ* for better explainability and to impose weak regularizations on relevant parameters, a truncated normal prior on the slope coefficient *k* to ensure positivity, and inverse-gamma priors for the scale hyper-parameters *σ*, which tend to drive parameters further away from zero than (truncated) normal priors.

### Model estimation and comparison methods

We performed Bayesian estimation with Stan^25^. See Supplementary Methods for details and Table S1 priors. For each dataset, the hierarchical model fits the data from all participants simultaneously. Note that the MAL measurements are non-evenly spaced. In DOSE, the smallest spacing is 1 week and in EXCITE, the smallest spacing is 2 weeks. To compare the hyper-parameters in both datasets, we used a time-step of 1 week for both datasets and considered non-available data as missing. The Bayesian method smoothly deals with the issue of missing data.

For both datasets, we compared the best model of Table S1 with models without the forgetting term, the learning term, or the self-training term in equation 1. We then compared the full model to a model without random effects (no between-subject variability). We finally compared the full model to “fixed” linear and logistic models. Model comparison (e.g., between the best model and simpler models) was first performed using the WAIC (Watanabe-Akaike information criterion), which is widely used to compare Bayesian models^26^. We also compared the models using leave-one-out cross-validation. See Supplementary Methods.

### Evaluation of forecasting accuracy on unseen data with the DOSE dataset

We then evaluated the long-term forecasting accuracy in four different scenarios on the DOSE dataset (see Results). We used a leave-one-subject-out procedure, which repeatedly re-fits the model while masking a number of last MAL measurements for a “left-out” participant. This procedure allows us to examine the model’s ability to utilize a database of “past” participants to predict the outcome of a “new” participant (with few data). We also compared the full model with a model without the hierarchical structure. We quantified the model’s forecasting accuracy using a modified RMSE, named Bayesian forecasting RMSE (BF-RMSE). The BF-RMSE is the RMSE between the forecasted values (one for each posterior draw) and a masked future measurement for a participant. We computed the mean BF-RMSE at a given future time-point by averaging over participants and one-sided *p*-values for the mean BF-RMSE using permutation tests.

## Results

### Fitting the hierarchical Bayesian state-space models

#### DOSE dataset

Figure 1A shows examples of fits to the DOSE data for the “best” hierarchical Bayesian model (see equations 1-3 in Methods) with the corresponding individual parameters (forgetting rates, learning rates, and self-training rates) for eight representative participants (two per dose). The model achieved good convergence, with R-hat < 1.05 for all parameters. The fit was overall excellent, with RMSE = 0.28 for all 40 participants (individual RMSE 0.26 ± 0.098), which is less than 6% of the MAL 0–5 scale. The median forgetting rate for each subject was between 0.83 and 0.89, in line with the mean estimated forgetting rate in our previous work with the EXCITE dataset of 0.86 ^12^. However, there was large between-subject variability. Whereas several participants showed a weak training effect with a median learning rate of less than 0.1 (e.g., D27), some showed learning rates above 0.4 (e.g., D23). Whereas a combination of small learning and self-learning rates is highly detrimental for long-term performance (e.g., D27, D37), the opposite yields improved long-term outcomes (e.g., D12).

**Figure 1:**
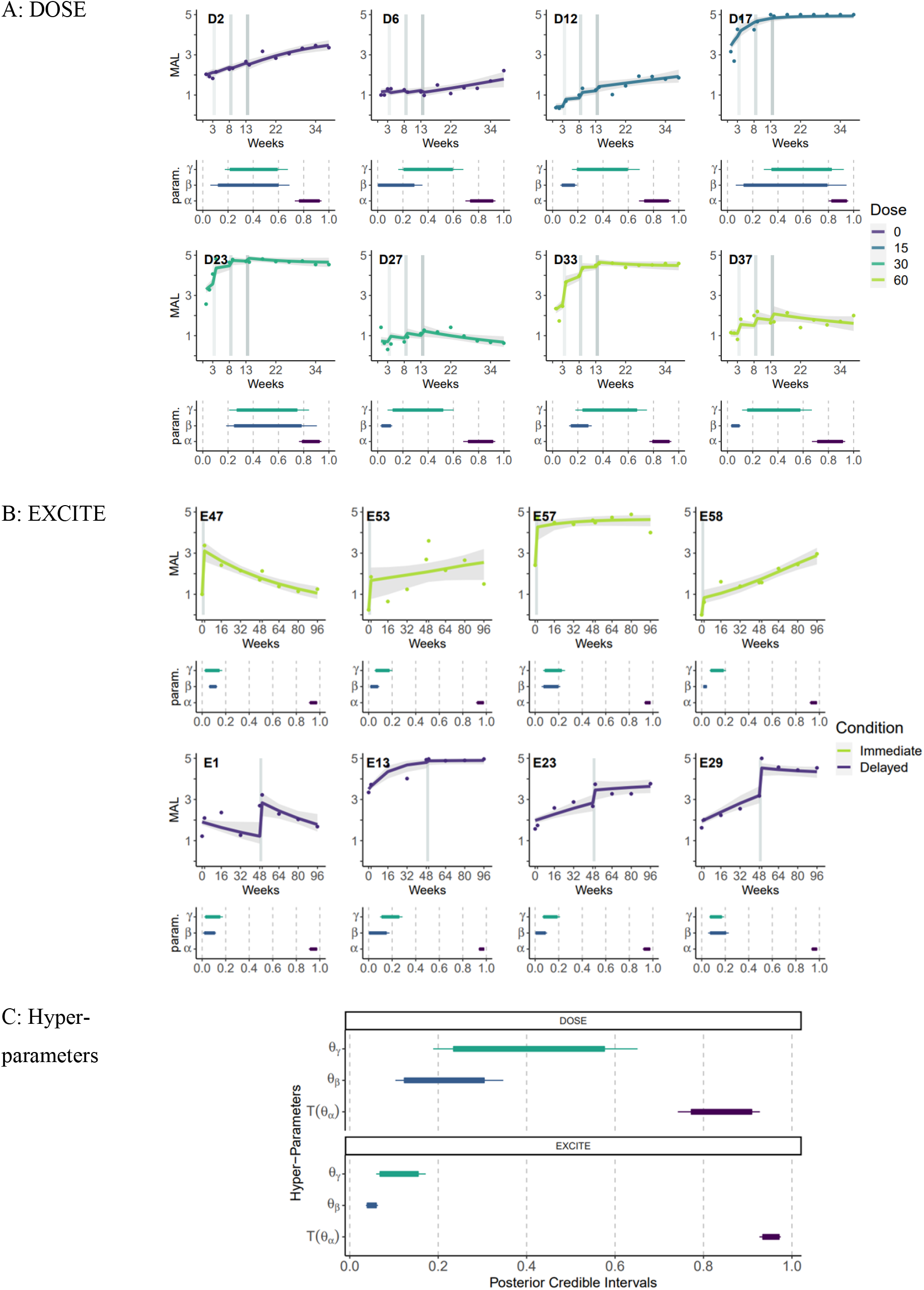
Data, model fit, and parameter estimates for the best learning model. A. DOSE data: Example of data, model fit, and parameter estimates for eight participants arranged by doses of training. B. EXCITE data. Example for eight participants arranged by the timing of training (immediate vs delayed). Upper panel for each participant: MAL data and fit for the best model. Dot: data. Lines: mean model fit. Shaded area: 95% CI. Lower panel: Posterior parameter distributions of the three main parameters (self-training rate γ_*i*_, learning rate *β*_*i*_, and forgetting rate α_*i*_). The thick bars show the 95% parameter CI and the thin bars the 99% CI. C. Hyper-parameters. Posterior distributions for the corresponding hyper-parameters. Note that T(θ_α_) is the transformed θ_α_ parameter (via the sigmoid function), which corresponds to the median of the logit-normal prior distribution for individual forgetting rates (α_*i*_).

We then verified that the motor learning model accounts for the phenomena previously observed in the DOSE study ^13^: immediate dose-response, the decrease in efficiency with both additional dose and weeks of training, the overall decay following training, the negative dose-dependent effect of training on retention, and the threshold above which self-training becomes sufficient to further increase the MAL without supervised training in the DOSE data. See Supplementary Results for details.

#### EXCITE dataset

Figure 1B shows examples of fits of the same “best” model to the EXCITE data and parameters for eight participants (four in each group: immediate and delayed). Here, again, the model achieved good convergence, with R-hat < 1.05 for all parameters. The mean RMSE was 0.35 (individual RMSE 0.32 ± 0.16), which is 7% of the MAL 0-5 scale. Thus, despite the few data points available per participants in EXCITE (9 instead of 14 in DOSE), the fit is still very good. Here again, there was large variability in the parameters between participants, notably for the effect of training. For instances, participants E47 and E1 showed pronounced decay outside of training, as captured by relatively small forgetting and self-training parameters. In contrast, participants whose MAL continues to increase post-training showed high self-training parameters. Finally, as for DOSE, training effects vary significantly between subjects (e.g., E57 and E29).

Comparison of the hyper-parameters between the two datasets (Figure 1C) shows that, as a group, the EXCITE participants have smaller learning rates (95% CI 0.040 – 0.06 for 0.12 -0.30), smaller self-learning rates (95% CI: 0.068 – 0.15 vs. for DOSE: 0.23 – 0.57), and larger forgetting rates (95% CI: 0.93-0.97, corresponding to a time constant of decay 15.2 – 34.6 weeks, vs. for DOSE: 0.77-0.90, time constant 3.4 – 11.0 weeks). We comment on the possible reasons for these differences in the Discussion.

#### Comparison with simpler models

The model comparison results show strong evidence that all three main terms in the state-space equation, forgetting, learning, and self-training, are required for a good fit for both DOSE and EXCITE datasets. Omitting the learning term has the biggest effect in worsening the fit in both DOSE and EXICTE, followed by the forgetting term and the self-training term. Not surprisingly, given the diversity of the MAL trajectories, the model with fixed effect only (i.e., for which the parameters are not individualized) performed the worst. For details, see Supplementary Table S2A for DOSE and Supplementary Table S2B for EXCITE.

Finally, we compared the model with two non-dynamical regression models: a linear model of time and a modified logistic model (see Methods) for both datasets. The logistic model performed relatively well compared to other models, such as the dynamical models without learning, but still largely worse than the full models and the dynamical models without self-training. This illustrates that “fixed” (non-dynamical) models, which cannot simultaneously account for the effects of learning and forgetting, respectively, cannot account well for our rehabilitation data.

#### Evaluation of forecasting accuracy on unseen data: Increasing accuracy and precision with additional outcome data

We then evaluated the long-term forecasting accuracy for new participants in four different realistic scenarios in which the clinician would assess and re-assess predictions as additional outcome data becomes available (see Methods). Figure 2A shows the hierarchical model’s forecasting fits (median, 95%, and 99% CIs) on four representative participants for the duration of the DOSE trial. The mean BF-RMSE (over participants) at six months post-training was 1.36 when only the baseline data were available, it then decreased to 0.91, 0.79, and 0.69 when the MAL data following the 1^st^, 2^nd^, and 3^rd^ bouts of training were available. Thus, when MAL data following the 2^nd^ bout of training is available, predictions at 6 months become remarkably accurate, in line with the Minimal Clinically Important Difference for the MAL of ∼0.5 ^27^.

**Figure 2:**
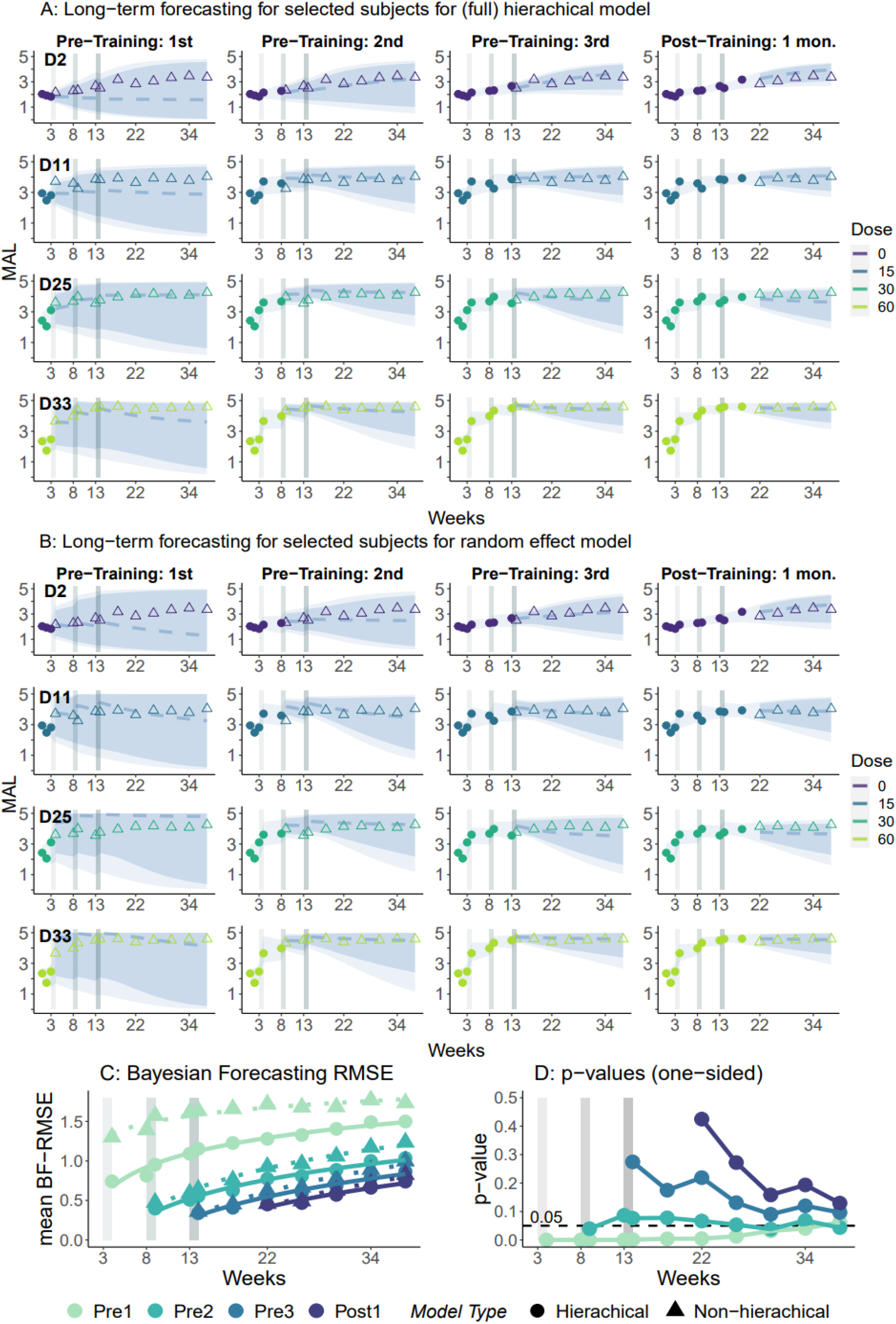
Accuracy of individual long-term forecasting **A. and B**. Examples of individual predictions for different amounts of incoming data with population priors for four participants, one per dose. D2: 0 hours. D11: 15 hours. D25: 30 hours. D33: 60 hours, each in four scenarios in which the availability of outcome measures increases for each participant (circles). B. Same as in A, but here, we do not use population hyper-priors but weakly-informative priors for each participant. Solid lines: model prediction (posterior median). Blue shaded zones (from darker to lighter): 90% and 95% prediction intervals. Triangles. MAL data measured but not used to update the model. **C**. Comparison of mean (Bayesian) RMSEs for each week in the different forecasting scenarios. Dots and Triangles are the mean RMSEs for the hierarchical and random effect (i.e., non-hierarchical) models, respectively. Solid and dotted lines are the best fit lines (log-linear) of the weekly mean RMSEs for the hierarchical and random effect models, respectively. **D**. One-sided p-values for the difference in mean RMSEs between the hierarchical and the random-effect models for each forecasting scenario.

Finally, we find that the random-effect model (when compared with hierarchical model) was largely inaccurate when only the baseline data were included (see Figure 2C and p-values between the two models in Figure 2D). Whereas the benefit of the hierarchical structure is evident when fewer data are available. Indeed, after the 3^rd^ bout of training, predictions with and without the hierarchy become nearly indistinguishable, as can be observed by comparing the individual plots in Figures 2A-B, and 2C. In this case, the *p*-values of the differences between the two models are high, notably for short-range forecasts (Figure 2D). However, interestingly, for long-range forecasts the p-values decrease again for all scenarios, showing the relative advantage of the hierarchical model (although not reaching the 5% significant level). This occurs because as percentage of missing data increases in long-range forecasts, information sharing across participants becomes once again more advantageous.

## Discussion

Our study presents a novel model to forecast a continuous motor outcome following rehabilitation in chronic stroke survivors. Although previous “static” models can predict the response to interventions using baseline data, these models cannot account for the changes in the outcome due to training, self-training, and forgetting in the long-term. Here, we showed that the accuracy of the prediction is much improved by a dynamical forecasting model that incorporates initial responses to training. Our results using data from both the DOSE and EXCITE clinical trials in chronic stroke showed that the best model is a non-linear state-space model derived from our previous research ^12,13^ that contains a forgetting term that depends on the memory at the previous time-step, a learning term as a function of the dose of supervised training, and the self-training term that uses the predicted MAL fed back to the memory model. Despite its relative simplicity, our motor learning model clearly captures the dynamics of the MAL in response to and following training in both datasets. In addition, it accounts for a number of phenomena previously observed in the DOSE study ^13^ (see Supplementary Results).

Comparisons of the hyper-parameters between the two clinical trials show smaller learning rates and self-learning rates and forgetting rates closer to 1 (corresponding to less decay) in EXCITE. The smaller learning rates can be explained by our previous study showing that the gains due to therapy^13^ (see also Supplementary Materials) decrease with larger doses. Since all participants in EXCITE received 60 hours of therapy, compared to an average of 23.5 hours of nominal therapy for DOSE, it is expected that the learning rates for DOSE will be greater. Another possibility is that the type of therapy in DOSE (ASAP) is more effective than that of EXCITE (constraint induced movement therapy), but a larger group of 60 hours ASAP training would be needed to test this hypothesis. Similarly, greater self-learning hyper-parameter for the DOSE group is consistent with our previous findings that self-learning decreases with the dose of therapy^13^. Finally, in our previous study, we also showed that the greater the gains due to therapy, the greater the forgetting^13^. Thus, a possibility is that constraint-induced movement therapy training in EXCITE is less effective than ASAP in DOSE, leading to less forgetting. A larger study with diverse range of time since stroke (i.e., participants in the acute, sub-acute, and chronic stages), doses, and types of therapy is needed to test these possibilities.

Although hierarchical Bayesian state-space models have not yet been used to inform the practice of neurorehabilitation, hierarchical Bayesian modeling has recently been used to model “spontaneous recovery” in the acute and pos-acute phases post-stroke across multiple clinical sites^1^. In our model, the hierarchical structure improved predictions both early in training and for long-term forecasts for a given patient compared to a non-hierarchical model (see Figure 2A) by “borrowing” information between patients. New incoming data, as well as prior knowledge, can be naturally incorporated in our model, yielding an on-line supervised learning method that continuously improves the predictions. Importantly, in the hierarchical models, learning occurs simultaneously at the individual and population levels: additional data improves the forecast for the current participant and the population overall. In future work, a higher-level site hierarchy can be added when models fit data from multiple sites, as in ^1^.

In future applications, hierarchical Bayesian state-space models can be used in “precision rehabilitation.” Currently, the dose and schedule of rehabilitation are determined based on clinical setting (e.g., in-patient, out-patient), historical precedent, and results from clinical trials. However, with an accurate forecasting model, one can determine optimal schedules of motor training that maximize expected outcomes. For instance, in previous work in motor learning ^28^, we used optimal control theory to determine the schedule that maximizes the mean long-term performance predicted by a non-Bayesian motor learning model. Such predictions of the mean are useful in data-rich applications. However, in personalized medicine, and especially in neurorehabilitation, the data are sparse, and the uncertainty of the predicted outcomes is high. In contrast, the current hierarchical Bayesian forecasting model generates a full distribution of the long-term outcomes based on the parameter uncertainty and measurement noise. This allows us to visualize both accuracy and precision of the model predictions with ease.

In contrast to our theory-driven approach, one can envision a purely data-driven approach to forecasting motor outcomes post-stroke using “black box” models, e.g., ^29^. Although such models may yield good or even better accuracy, they are not interpretable as they contain numerous parameters. In contrast, our approach yields an understanding of the mechanism(s) underlying the predictions and therefore an understanding of causes for individual differences in forecasts -see also for a related (non-Bayesian) example ^30^. Analysis of the model parameters can help the clinician make informed decisions about therapy ^5^. For instance, if a patient has a large learning rate but also a large forgetting rate, then it is predicted that the gains due to additional therapy will be short-lived. Or if the learning rate is near zero, then it can be predicted that even large doses of therapy will not yield large gains.

Notwithstanding, there are two main limitations in our study. The first limitation is the use of the MAL which relies on self-reported ratings of the quality of movement across a range of tasks. Nonetheless, it is striking that our simple model of motor learning well characterizes the dynamics in the MAL outcome in the two datasets, since the MAL score results from an average of scores for 28 activities of daily living. Relatedly, our “motor memory” variable cannot be taken literally as a neural variable, but rather as an aggregate of the multiple motor memories needed to perform the MAL. A second limitation is the limited size and relative homogeneity (because of the restricted entry criteria) of the datasets. As a result, a single covariate was included in the model, the initial MAL. Biomarkers derived from transcranial magnetic resonance and brain imaging would allow us to refine predictions, notably soon after stroke when the predictions are poor, in a more diverse population. In addition, the predictions would further improve with a larger number of measurements from each individual. Further, clinical studies that use connected objects and sensors, e.g., ^31^ would allow the collection of such datasets that, together with a forecasting model such as that proposed here, would form the basis of precision rehabilitation for not only arm and hand function post-stroke but also other functions, such as gait rehabilitation post-stroke, or neurologic conditions such as traumatic brain injury or spinal cord injury.

## Data Availability

All data produced in the present study are available upon reasonable request to the authors

## Declaration of Conflicting Interests’ Statement

The authors declare that there is no conflict of interest related to this work.

## Funding

This work was supported by NIH grants R56 NS100528 and R21NS120274 to NS and P41-EB001978 and the Alfred E. Mann Institute at USC to DD.

## Supplementary Materials

### Supplementary Methods

#### Bayesian parameter estimation methods

Fitting the dynamical model given by equations 1-3 is performed via Bayesian inference, which incorporates the prior distributions and data to generate a posterior distribution for each random variable via the Bayes’ rule. We imposed weakly-informative priors on the hyper-priors. In particular, the priors on μ_α_ and *σ*_α_, were chosen such that the median of the posterior distribution for forgetting rates (of all participants) is 0.85, in line with the median forgetting rate estimated in our previous work (0.86) with the DOSE dataset ^13^. Similarly, the hyper-priors for the learning rate and self-training rate generate values in the ranges of the parameters found in our previous studies ^12^ with the EXCITE dataset and ^13^ with the DOSE dataset – see Table S1.

**Table S1.** Subject-dependent parameters and population hyper-priors used in the best model. Note that we modeled the measured 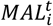with a generalized *t*-distribution centered at 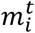with a scale parameter *σ*_*MAL*_ the degrees of freedom *v*.

| Model Parameters | Priors | Hyper-priors |
| --- | --- | --- |
| Forgetting rate | $\alpha_i \sim 1/(1 + e^{-\mathcal{N}(\theta_\alpha, \sigma_\beta)})$ | $\theta_\alpha \sim \mathcal{N}(2, 1)$<br>$\sigma_\alpha \sim \text{InvGamma}(3, 2)$ |
| Learning rate | $\beta_i \sim \mathcal{N}_{[0, \infty)}(\theta_\beta, \sigma_\beta)$ | $\theta_\beta \sim \mathcal{N}(0, 1)$<br>$\sigma_\beta \sim \text{InvGamma}(4, 2)$ |
| Self-training rate | $\gamma_i \sim \mathcal{N}_{[0, \infty)}(\theta_\gamma, \sigma_\gamma)$ | $\theta_\gamma \sim \mathcal{N}(0, 1)$<br>$\sigma_\gamma \sim \text{InvGamma}(4, 2)$ |
| Initial state of memory | $x_i^0 \sim \mathcal{N}_{[0, \infty)}(k \text{ MAL}_{\text{ini}}, \sigma_{\text{ini}})$ | $k \sim \mathcal{N}_{[0, \infty)}(0, 2)$<br>$\sigma_{\text{ini}} \sim \text{InvGamma}(3, 2)$ |
| Data | Likelihood | Hyper-priors |
| MAL (measured) | $\text{StudentT}(\text{MAL}_i^t \mid m_i^t, \sigma_{\text{MAL}}, \nu)$ | $\nu \sim \text{Gamma}(2, 0.1)$<br>$\sigma_{\text{MAL}} \sim \mathcal{N}_{[0, \infty)}(0.25, 0.1)$ |

We used 6 chains for 10000 iterations, with 5000 warm-up samples. No additional iterations were needed, as all parameters attained the between-chains Gelman-Rubin convergence statistic *r-hat* < 1.1, as recommended. We then used *tidybaye* ^32^, *MCMCvis* and *ggmcmc* packages to generate summary statistics and plots of the posterior distributions for the parameters and variables and our model. A modified leave-one-out cross-validation (see below) was performed to evaluate the model’s ability to forecast individual future outcomes using the supercomputer at the Center for Advanced Research Computing (CARC) of the USC. After fitting the model, we report 95 % and 99% posterior credible intervals for the individual parameters, the population parameters, and the predicted MAL at each week.

#### Model comparison methods

For both datasets, we compared the best model (described above and in Table S1) with several reduced models. We first compared simpler models without either the forgetting term, the learning term, or the self-training term to the complete model of equation 1. We also compared the full model to a model without random effects (no between-subject variability). We finally compared the full model to “fixed” linear and logistic models, in which the state-space equation (1) is replaced by a simple linear function of weeks of the form:

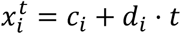

where *c*_*i*_, *d*_*i*_ are arbitrary coefficients (for subject *i)* and *t* is the number of weeks after the initial MAL measurement. The logistic model, but not the linear model, contained the modified sigmoid function (equation 2) that converts motor memory into predicted MAL.

Model comparison (e.g., between the best model and simpler models) was first performed using the WAIC (Watanabe-Akaike information criterion), which is widely used to compare Bayesian models ^26^. The WAIC is an estimation of expected log point-wise predictive density (ELPD) that adjusts for overfitting using the effective number of parameters. The ELPD is a theoretical/ideal measure of a model’s predictive accuracy ^26^. Maximizing ELPD is equivalent to minimizing the KL-divergence of the true data-generating process to the posterior predictive distribution.

We also compared the models using the *elpd_loo*, which is the ELPD approximated using Leave-One-Out Cross-Validation (LOO-CV), which allows us to compute the *elpd_diff*, a measure of the difference in the *elpd_loo* between two models as well as the 95% confidence intervals of difference. The *elpd_loo* has shown to be asymptotically equal to WAIC but also “more robust in the finite case with weak priors or influential observations” ^26^. Note that when comparing several models, the *elpd_diff* is only computed for each model against that of the best model (which has the largest *elpd_loo*). Then, under the normality assumption, we can estimate *Pr*(*better*), the probability that to-be-compared model is better than the best model found through PSIS-LOO (i.e., when the true *elpd_diff* ≥ 0).

## Supplementary Results

### Model comparison

**Table S2A:** Model comparison for DOSE. *WAIC*: Watanabe-Akaike information criterion. The models are ordered from best to worse, as ranked by the *WAIC. Elpd*: expected log point-wise predictive density. *Elpd_loo*: the ELPD approximated using Leave-One-Out Cross-Validation (LOO-CV); *elpd_diff*, a measure of the difference in the elpd_loo between two models. *se_diff* is the (estimated) standard error of *elpd_diff*. The false-positive rate is the probability that the best model is worse than the to-be-compared model. See Methods for details.

|  | *WAIC* | *elpd_loo* | *elpd_diff* | *se_diff* | *95% CI* | *Pr(better)* |
| --- | --- | --- | --- | --- | --- | --- |
| ***Full (best)*** | 296.74 | -149.42 |  |  |  |  |
| ***No self-training*** | 346.35 | -173.76 | -24.35 | 10.14 | (-44.22, -4.47) | 8.19E-03 |
| ***Logistic*** | 487.55 | -244.83 | -95.41 | 17.79 | (-130.27, -60.55) | 4.07E-08 |
| ***No forgetting*** | 496.91 | -249.50 | -100.08 | 17.54 | (-134.46, -65.71) | 5.77E-09 |
| ***No learning*** | 506.92 | -253.95 | -104.54 | 15.14 | (-134.21, -74.86) | 2.51E-12 |
| ***Linear*** | 512.34 | -256.55 | -107.13 | 18.60 | (-143.58, -70.68) | 4.20E-09 |
| ***Fixed Effect*** | 588.35 | -294.26 | -144.85 | 19.77 | (-183.6, -106.09) | 1.19E-13 |

**Table S2B:** Model comparison for EXCITE. See Table 2A for abbreviation.

|  | *WAIC* | *elpd_loo* | *elpd_diff* | *se_diff* | *95% CI* | *Pr(better)* |
| --- | --- | --- | --- | --- | --- | --- |
| ***Full (best)*** | 914.58 | -458.13 |  |  |  |  |
| ***No forgetting*** | 1079.25 | -539.62 | -81.50 | 19.01 | (-118.76, -44.23) | 9.08E-06 |
| ***No self-training*** | 1181.18 | -591.06 | -132.93 | 20.81 | (-173.73, -92.13) | 8.49E-11 |
| ***Logistic*** | 1357.74 | -680.91 | -222.78 | 27.34 | (-276.36, -169.21) | 2.22E-16 |
| ***Linear*** | 1393.45 | -698.18 | -240.05 | 28.04 | (-295.01, -185.09) | 0.00E+00 |
| ***No learning*** | 1400.08 | -701.61 | -243.49 | 26.89 | (-296.19, -190.79) | 0.00E+00 |
| ***Fixed Effect*** | 1501.13 | -750.99 | -292.86 | 27.70 | (-347.16, -238.56) | 0.00E+00 |

### Qualitative model evaluation

Using the best model described above, we tested whether the best motor learning model could, despite its simplicity, reproduce previous findings of the DOSE trial^13^: an immediate dose-response, a decrease in efficiency with both additional dose and weeks of training, an overall decay following training, a negative dose-dependent effect of training on retention, and the threshold above which self-training becomes sufficient to further improve the MAL without supervised training. For this, we simulated the model of equations 1-3 using all posterior samples of the individual parameters for each of the 40 participants in the DOSE datasets. We processed and plotted the predicted MAL data in a similar fashion to our previous study ^13^: i) To compute the short-term efficacy, we computed the changes in MAL due to the three bouts of training. ii) To compute the dose efficiency, we divided the changes in MAL due to the three bouts of training by each dose. iii) To compute the decay following training for each dose, we computed the weekly change in MAL in two-month intervals following training divided by the number of weeks in each interval. iv) Finally, to determine the threshold above which self-training can further increase outcomes, we fitted a regression model of the weekly changes in MAL from immediately post-training to 6 months post-training as a function of the mean MAL post-training with dose as a factor.

The model reproduced our six previous results. First, as shown in Figure S1A, the model replicated the increase in the efficacy of training with greater doses, with a near-linear dose-response relationship. Second and third, the efficiency of training depends on both the weekly dose and the number of training bouts (Figure S1B). Increasing the number of hours of training resulted in a decrease in efficiency in an exponential-like fashion. Similarly, across dosages, the first bout of training increased the MAL, whereas the additional bouts were increasingly less efficient (Figure 1B). Fourth and fifth, retention depends on time post-stroke and the dosage of training. For the smallest dose, the MAL increases following training (positive Δ*MAL*/*week*), with the initial increase larger than in later months. In contrast, for large dosages, the MAL decreases (negative Δ*MAL*/*week*), with the initial decay larger than the decay in later months (Figure S1C). Whereas such an exponential-like convergence is not surprising given our choice of a first-order state-space model (see examples of fit for 0- and 60-hours dose in Figure 1), these results match those of our previous study in which we approximated the retention data with three linear segments of two months each^13^. Sixth, these opposite retention results for small and large doses are due to the positive “self-training” effect counteracting the negative effect of increasing dose (the greater the dosage, the more the forgetting), as shown in Figure S1D. When the MAL post-training is above a dose-dependent threshold, the MAL keeps increasing, as we previously found with the piece-wise model^13^.

**Figure S1:**
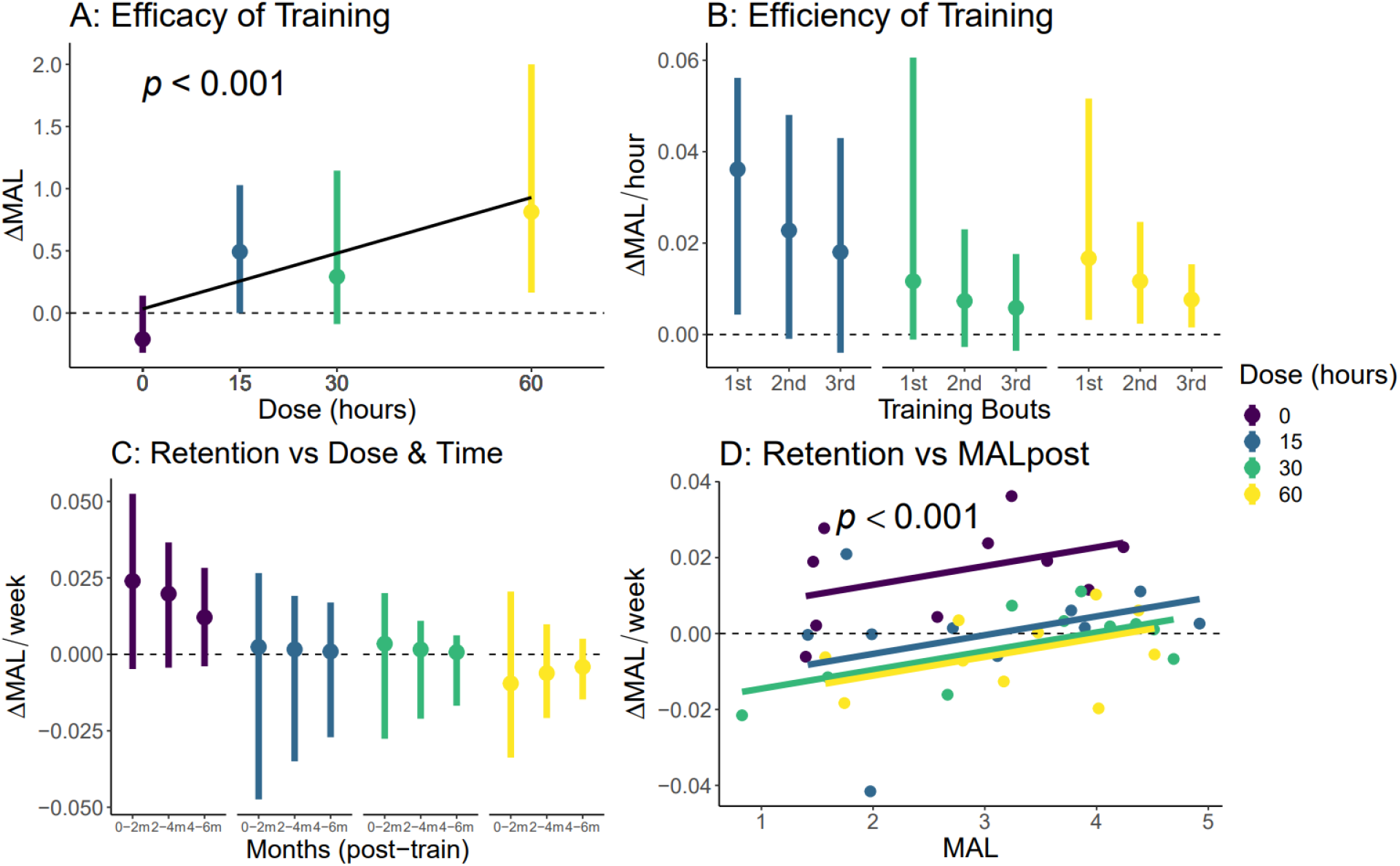
Qualitative validation of the best motor learning model. **A**. Dose efficacy: Effect of dosage on the MAL during supervised training. **B**. Efficiency of training for different training dosages and for the three bouts. Efficiency is computed by the gain in MAL per hour of training.**C**. Effect of time post-training on retention for each dosage: Shown are the forgetting rates in the six months following supervised training as a function of months post-training. **D**. Effect of average post-training MAL on the gain in MAL. The colored lines show retention as a function of average post-training MAL for different dosages, and the colored dots show the individual retention rates. In C and D, the dotted line indicates the retention rate of 0, indicating no change. In A, B, and C, the dots are the population means based on predicted MALs, and the bars the 95% CI.

## Footnotes

a For instance, a (hypothetical) treatment A, which is predicted to increase the outcome by 50% with a 95% confidence interval of 5%, may be preferred to treatment B, which is predicted to increase the outcome by 60% with a 95% confidence interval of 20%.

## Notes

### Competing Interest Statement

The authors have declared no competing interest.

### Clinical Trial

DOSE clinical trial ID NCT01749358. EXCITE clinical trial ID NCT00057018

### Funding Statement

This work was funded by NIH grants R56 NS100528 and R21NS120274 to NS and P41-EB001978 and the Alfred E. Mann Institute at USC to DD.

### Author Declarations

The Institutional Review Board of the Universty of Southern Californial gave ethical approval for the DOSE clinical trial. The Institutional Review Boards of the following seven participating sites gave ethical approvals for the EXCITE clinical trial: Emory University (Georgia) The Ohio State University (Ohio) University of Alabama at Birmingham (Alabama) University of Florida at Gainsville (Florida) University of Southern California (California) University of North Carolina at Chapel Hill (North Carolina) Wake Forest University (North Carolina)

